# Evaluating the GRACE voice assistant for dementia care among caregivers and healthcare professionals: An interview study

**DOI:** 10.1101/2025.10.08.25337586

**Authors:** Florianne Walliser, Nikola Biller-Andorno, Tobias Kowatsch, Rasita Vinay

**Affiliations:** School of Medicine, University of St. Gallen, St. Gallen, Switzerland; Institute of Biomedical Ethics and History of Medicine, University of Zurich, Zurich, Switzerland; Institute for Implementation Science in Health Care, University of Zurich, Zurich, Switzerland; Centre for Digital Health Interventions, Department of Management, Technology, and Economics, ETH Zurich, Zurich, Switzerland

**Keywords:** Dementia, people with dementia, digital health intervention, digital assistive technology, voice assistant, dementia care, informal caregivers, healthcare professionals, user acceptance, human-robot-interaction, qualitative interview study

## Abstract

**Background:** Considering rising dementia rates, voice assistants are being investigated to support people with dementia (PWD) and reduce the burden on caregivers (CGs). GRACE, an intervention-delivering voice assistant, provides cognitive support and assistance with daily living for people with early-stage dementia. Initial pilot trials showed promising user experience among healthy adults but required further evaluation by care professionals working with PWD as a next reiteration process.

**Objective:** This study investigated three main research questions: (1) Which intervention components of GRACE do healthcare professionals (HCPs) and CGs find most valuable for supporting the daily lives of PWD? (2) What key constraints do HCPs and CGs expect when implementing GRACE in dementia care settings? and (3) What changes or additions do HCPs and CGs recommend for optimizing the functionality of GRACE?

**Methods:** Twelve semi-structured interviews were conducted (CG: n=4, HCP: n=8). Anonymized transcripts were coded and analyzed thematically to directly answer the research questions.

**Results:** Respondents found the onboarding activity and memory exercise to be most valuable, due to its relevance in everyday life. Identified barriers for implementation included practical, technical and ethical challenges. Recommendations for successful implementation and further development of GRACE included improving PWD acceptance and greater AI-supported personalized content. Finally, a list of future exercises for incorporation and other use cases of GRACE beyond dementia care were also identified.

**Conclusions:** GRACE has great potential as a complementary tool in dementia care. Ensuring personalization, ease of use, and alignment with care processes will be critical for adoption and long-term success.

## Introduction

### Background

In recent years, we have seen a rise of digital assistive technologies being used in dementia care with promising potential to support people with dementia (PWD) [1–4]. As the prevalence of dementia continues to grow, it intensifies the demand for individualized and specialized care, placing growing pressure on formal and informal caregiving structures [5]. Dementia contributes significantly to disability and dependency among older adults, presenting complex challenges not only for PWD, but also for healthcare professionals (HCPs), formal or informal caregivers (CGs) who provide daily support [4].

The World Health Organization projects a need for approximately 40 million new health and social care jobs by 2030 to meet the growing demand for dementia-related services [6]. Unlike other medical conditions, dementia care often requires highly specialized, personalized and adaptive approaches due to the variability of cognitive and functional symptoms across individuals [7–8]. This complexity is compounded by a global shortage of trained professionals [9], as well as family members (who would better understand the needs of PWD) being increasingly less available to provide full-time care.

In this context, digital assistive technologies such as voice assistants are being explored as tools to help improve quality of life for PWD while easing caregiving responsibilities [4]. Speech-enabled systems such as smart speakers show potential in supporting daily routines, promoting independence and offering cognitive stimulation [1, 10]. Despite these developments, there remains a lack of in-depth empirical research examining how such technologies are perceived in terms of acceptance, usability and usefulness by those directly involved in dementia care and how they can be meaningfully integrated into real-world care practices [11].

### The GRACE voice assistant

One such technology is the GRACE embodied voice assistant, designed to provide lifestyle and cognitive interventions to PWD (early stage) [12]. GRACE aims to go beyond personal assistive robots, and enable individuals to continue aging in place, thus impacting their overall quality of life while also addressing the CG shortage. The current version of GRACE provides various voice-based exercises, which individuals can use in their homes at their own pace. Currently, GRACE has been pilot-tested and reiterated among healthy adults (first with 18-60 years old, then with older adults aged 60 or above), to understand initial technological feasibility and perspectives on its provided interventions [13–15]. The interventions were developed according to Cognitive Stimulation Therapy (CST), an evidence-based non-pharmacological intervention for mild to moderate dementia [16], and delivered through a natural spoken interface with a digital coach (GRACE). Results from these pilot studies showed that users found GRACE to be easy to use, useful, considerate of ethical challenges, and able to form a working alliance (i.e., a collaborative relationship between the user and GRACE) [14–15]. Future studies of GRACE aim to further iterate the technology, including its hardware, infrastructure and design more interventions informed by CST, through focus group studies and pilot studies among PWD. After final iterations, GRACE would be evaluated in a feasibility study with PWD and their CGs at home [17].

### Objectives

The aim of this study is to evaluate the current GRACE prototype (version 2) with respect to its perceived usefulness, usability, and integration potential into dementia care, as viewed by healthcare professionals (HCPs) and caregivers (CGs). Specifically, the study investigates how these stakeholders value different intervention components, anticipate constraints to implementation, and also envision improvements. PWD were not included as part of this interview study, as we hope to slowly move towards the target population, by completing necessary reiterations of the technology, before finally engaging PWD. Accordingly, the following research questions are addressed:

**RQ1.** Which intervention components of GRACE do HCPs and CGs find most valuable for supporting the daily lives of PWD?
**RQ2.** What key constraints do HCPs and CGs expect when implementing GRACE in dementia care settings?
**RQ3.** What changes or additions do HCPs and CGs recommend for optimizing the functionality of GRACE?

## Materials and Methods

### Study design

We conducted a qualitative semi-structured interview among participants in the German-speaking region of Switzerland and Germany, to evaluate their perspectives on the GRACE voice assistant. The study was conducted from October 2024 to May 2025. The ethics committee of the University of St. Gallen reviewed the study protocol and checklist and provided an exemption from a formal review on 17 September 2024.

### Study participants

Participants were recruited through multiple dimensions: via a study flyer, online dissemination on the study website, institute websites, and LinkedIn, as well as via emails to local organizations and personal networks of the researchers that were identified to be relevant in dementia care. Participants included both CGs who provided care to PWD and HCPs working within dementia care. Individuals were eligible to participate in the study if they: (1) could speak and understand German, (2) professionally or privately care/support PWD, and (3) were able to participate online. All participants received a detailed study information sheet and provided signed informed consent prior to the scheduled interview.

### Data collection

The interviews were conducted in German, and took place online via Microsoft Teams, however one interview was in-person and another via telephone due to technical difficulties. All participants provided consent for recording of the interview. An interview guide was developed by all authors, validated by an external expert, and was divided into profiling, content, and concluding sections. Participants were asked questions relating to the use of GRACE, its implementation and optimization. An interview protocol was completed by FW following each interview, reporting any contextual factors, such as disruptions or interview flow.

### Data analysis

The semi-structured interviews were audio-recorded and transcribed verbatim. All transcripts were anonymized and replaced with participant codes (i.e., CG# for caregivers and HCP# for healthcare professionals). All recordings were stored on a secure University of Zurich server, available only to the research team and deleted after transcription. Transcription was conducted using noScribe (version 0.5.0b) and manually reviewed by FW to ensure correct translation of Swiss German terms. Data was analyzed using inductive thematic analysis [18]. Transcripts and protocols were coded using ATLAS.ti (version 25.0.1), where initial codes were first generated, then aggregated into code groups relating to the study’s research questions and further categorized into sub-themes. The final codebook was reviewed by FW and RV.

## Results

A total of 12 semi-structured interviews were conducted for this study: 4 interviews with CGs and 8 interviews with HCPs who support PWD in their everyday lives. Table 1 displays the characteristics of the study participants. CG4 was conducted with two CGs who wished to participate together. To obtain specific feedback on the current performance and functionality of GRACE, all respondents were sent a short demo video prior to the interview (available on our OSF repository: https://osf.io/qtxkc). The video showed an interaction between GRACE and a user, from a previous pilot study. The interaction shown in the video consisted of four parts: 1) an onboarding section in which GRACE and the user briefly introduced themselves; 2) a guessing game exercise in which the user had to match animal sounds to the corresponding animals; 3) a memory exercise in which the user had to remember items on a shopping list and then repeat them; and 4) a final breathing exercise, which GRACE guided the user through. The interviews were then conducted based on this preparatory video.

**Table 1.** Participant characteristics for this interview study.

| Participant Code | Role | Intensity of contact with PWD | Experience using technology in care | Previously known to the interviewer |
| --- | --- | --- | --- | --- |
| CG 1 | Relative of PWD | high | none-low | yes |
| CG 2 | Relative of PWD | high | high | yes |
| CG 3 | Relative of PWD | medium | none-low | no |
| CG 4 | Relatives of PWD (dual interview) | high | none-low | yes |
| HCP 1 | Professional care worker | high | none-low | distantly |
| HCP 2 | Former therapist | high | none-low | no |
| HCP 3 | Professional care worker | high | medium | distantly |
| HCP 4 | Psychiatrist and psychotherapist | medium | medium | no |
| HCP 5 | Excluded in analysis due to poor quality |  |  |  |
| HCP 6 | Advanced Practice Nurse (APN) | medium | medium | no |
| HCP 7 | Nursing professional | high | medium | no |
| HCP 8 | Nursing professional | high | none-low | no |
| HCP 9 | Neuropsychologist | medium | medium | no |

*Note: CG – caregiver; PWD – person with dementia; HCP – healthcare professional. Intensity of contact with PWD was assigned as follows: low (approximately once per week or less), medium (several times per week), high (daily contact). Experience using technology in care was assigned as follows: none-low (no use of modern assistive technologies or only occasional use of long-established consumer devices such as radio), medium (use of technology for individual cases or specific purposes such as entertainment or monitoring purposes), high (regular and diverse use of technologies across multiple care activities). These levels were subjectively assigned by the interviewer following manual review of transcripts and post-interview protocols and are intended as descriptive indicators for sample characterization rather than as validated quantitative measures.*

In the first part of the interview, the respondents’ initial impressions of GRACE were gathered. All respondents had watched the GRACE video immediately before or during the interview (in the case of HCP1), enabling them to provide detailed feedback on the current GRACE exercises. Both the CGs and the HCPs were generally open and interested in GRACE. When asked directly, all respondents said they were interested in trying out GRACE themselves in the future and possibly using it. The general openness to use it was therefore very high. As for initial reactions to GRACE, the respondents expressed a mixture of appreciation, curiosity, skepticism, and constructive criticism. Many were impressed by the assistant’s friendly demeanor, especially in the onboarding, where GRACE introduces herself and asks a few personal questions. Participants found this initial interaction particularly valuable for reducing tension and building trust. In addition, initial reactions to the appearance of GRACE were mainly positive as well. Here, the visual design of GRACE was perceived as accessible and friendly by most respondents - an important factor when interacting with PWD according to the respondents.

### Evaluation of the onboarding activity

The onboarding activity, where GRACE briefly introduces itself and asks the user a few personal questions, was rated particularly positively by participants. Almost half the participants (n=5, 41.6%) said that this personal introduction made initial contact much easier and helped build trust between GRACE and the user. However, some of GRACE’s statements during the introduction were perceived as rather unnatural by the respondents. For example, when the user in the demonstration video answered that she had no siblings, GRACE replied with “I’m sorry.” Several respondents found this answer irritating, as it did not correspond to the social norms they were accustomed to. It was therefore noted that GRACE’s responses to such topics should be made even more natural in the future. Despite such constructive criticism, however, the onboarding was generally seen as a good starting point for interaction between GRACE and the conversation partner. Some of the respondents also noted that, ideally, this phase should be extended/expanded in the future to enable an even more natural, interesting and easy start to the interaction.

### Evaluation of GRACE’s exercises

Overall, the memory exercise received the best ratings. Five of the respondents even rated this exercise as the best so far. In contrast, only two respondents named the breathing exercise and none of the respondents named the guessing exercise as the best exercise. To better understand these assessments, detailed questions were then asked about each exercise to gather both positive and critical feedback on the current functionalities of GRACE. This feedback is summarized below for each exercise, noting that respondents focused their comments on the exercises they had the most to say about. Furthermore, it should be noted that at no point were respondents asked to name the best and worst exercises; they were simply asked where they saw the greatest potential for GRACE to support PWD, CGs, and HCPs in everyday life. This part was therefore designed to be relatively open-ended to capture the honest opinions of respondents as realistically as possible.

#### Memory exercise

Table 2 summarizes the feedback received relating to the memory exercise intervention component. Participants provided both positive and negative feedback, as well as suggestions for further improvement to the current memory exercise.

**Table 2.** Feedback of the memory exercise delivered by GRACE.

| Feedback | Mentioned by |
| --- | --- |
| *Positive feedback* |  |
| Rated as the best exercise. | CG2, CG3, CG4, HCP2, HCP8 |
| Appreciated its ability to activate and cognitively stimulate PWD. | CG2, CG3, CG4 |
| Thematic relevance for everyday life of PWD. | CG1, HCP1, HCP9 |
| Shopping list theme was praised for its simplicity and suitability for everyday use. | CG1, HCP1, HCP9 |
| Appreciated increasing level of difficulty over time. | HCP9 |
| **Negative feedback** |  |
| Found the level of difficulty too high. | HCP3, HCP7 |
| Criticized the corrective feedback from GRACE when the user forgot an item on the list, finding it harsh, and potentially causing discomfort to the PWD, or make them feel like they were being tested. | HCP3, HCP7, HCP9 |
| **Recommendations for improvement** |  |
| Replacing closed-ended questions with more open-ended questions. | CG2, CG3, HCP3, HCP4, HCP9 |
| Exchanging direct corrections with hints. | HCP9 |
| The topic (shopping list) could be expanding and personalized according to individual interests of the PWD. Topic recommendations included seasonal topics (Christmas, Easter) or personal hobbies (travel, history, geography). | HCP4, HCP7, HCP9 |
| Incorporating more praise. | HCP4, HCP9 |
| Simplifying and shortening instructions. | HCP7 |
| Communicating the progress of PWD over time. | CG1 |

Direct participant quotes were translated by the authors using DeepL, and some have been selected to represent the feedback summarized in Table 1.

> *“I also like the memory task with the shopping list. It’s something that’s very relevant to them, something that’s very close to them. (..) I can imagine that they will respond well to it because they feel that they can do it and want to do it themselves.”* (HCP9)
>
> *“I think (…) the memory exercise is quite good. But I have to say (…) that there are people (…) who don’t learn anything because it’s like a test. I hear this from our patients too when you do these tests, these memory tests, or when you do exercises where you have to repeat a word or remember what the other person said, or something like that. (…) Some people really find it unpleasant because it’s like an exam, like a test where they also realize that they don’t do so well there.”* (HCP7)

Regarding recommendations for improvement to the memory exercise, participants suggested that GRACE could instead ask first “What do you need for breakfast?” or “What usually goes on a shopping list?”. In terms of the feedback provided for missing recall items, it was also suggested for GRACE to instead provide hints such as “The item that is still missing is spread on bread in the morning” instead of “You forgot the butter”.

#### Breathing exercise

Table 3 summarizes the feedback received for the slow-paced breathing exercise, with an immersive environment. Participants shared a mix of positive and negative feedback, along with recommendations for enhancing the current breathing exercise.

**Table 3.** Feedback of the breathing exercise delivered by GRACE.

| Feedback | Mentioned by |
| --- | --- |
| **Positive feedback** |  |
| Rated as the best exercise. | HCP3, HCP7 |
| Found it good as an initial exercise (e.g. for its potential to relieve initial tension). | HCP2 |
| Instructions are easy to follow. | HCP7 |
| Useful when the PWD is open to participation. | CG3, CG4, HCP9 |
| **Negative feedback** |  |
| Effectiveness greatly depends on the openness of the PWD to this type of exercise. | HCP4, HCP8 |
| Questioned the added value of the exercise. | HCP6 |
| **Recommendations for improvement** |  |
| Use of Familiar Voices in Breathing Guidance | HCP7 |
| Simplification of Breathing Instructions | HCP1 |

A selected direct quote relating to this exercise, which nicely summarizes the feedback from Table 3 is:

> *“I personally think the breathing exercises are great. I’m a fan of these exercises. My mother would probably find it completely silly. Why should I breathe now? It’s very difficult. It probably really depends on the patient. But it’s a good idea. Anything that activates makes sense. You just have to try a lot and see what works and what doesn’t.”* (CG3)

#### Guessing exercise

Similar to the memory and breathing exercises, Table 4 summarizes the feedback obtained for the animal guessing exercise. Participants provided both positive and negative feedback, as well as recommendations for improvement.

**Table 4.** Feedback of the animal guessing exercise delivered by GRACE.

| Feedback | Mentioned by |
| --- | --- |
| **Positive feedback** |  |
| Found it useful for PWD. | HCP4, HCP6, HCP9 |
| Found the simplicity as particularly positive. | HCP4 |
| **Negative feedback** |  |
| Found it too childish. | CG1, CG3 |
| Questioned its overall usefulness. | HCP8 |
| **Recommendations for improvement** |  |
| To expand the range of topics and sounds to reflect the interests and life experiences of PWD to make it more interesting and fun. | CG1, CG2, HCP1, HCP4, HCP7, HCP9 |
| More praise and positively framed feedback on the PWD's progress should be incorporated. | CG1, HCP2 |

Additionally, some participants also mentioned the involvement of family members or CGs in this exercise as a way to provide/improve personalization. One participant also imagined scenarios in which former farmers could hear milking sounds via GRACE, however also noted the technical complexity:

> *“There’s a lot of potential there. But that would probably require a lot (…) of development”* (HCP7)

### General impressions of GRACE

In addition to feedback on specific exercises, other general impressions of GRACE’s current appearance and functions were collected, these have been summarized in Table 5.

**Table 5.**
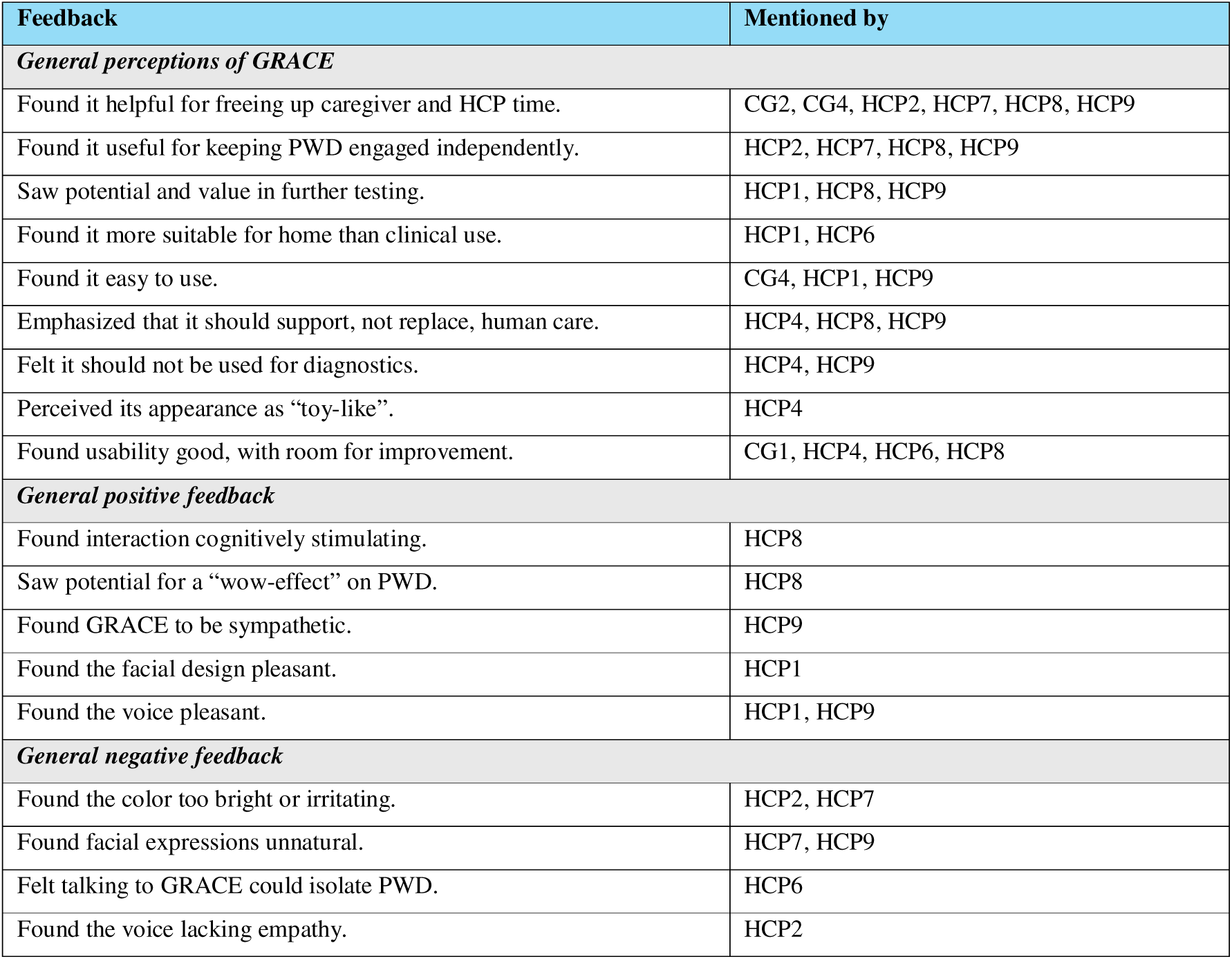
Feedback on the general impressions of GRACE.

In summary, the evaluation of the current exercises provided detailed insights into the strengths, weaknesses, and optimization potential of GRACE, as a direct response to RQ1. The onboarding was highly praised for its trust-building and tension-reducing effect, but minor improvements such as optimizing GRACE’s unnatural responses were suggested. The memory exercise received the most positive overall ratings and was appreciated for its potential to stimulate the brain and its suitability for everyday use. However, the corrective feedback and its “test-like” atmosphere was criticized. Recommendations included replacing corrections with hints and making it more open-ended and better tailored to the interests of PWD. The breathing exercise was generally found to be helpful for reducing tension, but it was noted that its effectiveness strongly depends on the openness of PWD to use it, suggesting possible barriers to acceptance. The guessing game exercise was praised for its simplicity but also criticized as too childish. An important suggestion was to expand the content and adapt it to the individual interests of PWD. Beyond the individual exercises, general impressions of GRACE showed mixed reactions to its appearance: some praised the friendly design and potential to reduce the workload of caregivers, while others rated the appearance and voice negatively. Ultimately it was emphasized that GRACE should serve as a complementary tool for CGs and HCPs instead of a replacement.

### Potential barriers to using GRACE

In addition to initial impressions, the interviews also explored potential challenges and barriers to the practical introduction and use of GRACE, in order to answer RQ2 (key implementation constraints). The findings focused on the willingness of CGs, HCPs and the expected willingness of PWD to interact with GRACE, its integration into workflows, and existing ethical concerns among the respondents (see Table 6).

**Table 6.** General adoption and ethical challenges as reported by respondents.

| Feedback | Mentioned by |
| --- | --- |
| ***On the part of the PWD*** |  |
| Lack of motivation to use GRACE. | CG1, CG2, CG3, CG4, HCP1, HCP3, HCP7, HCP8, HCP9 |
| Limited technological familiarity or openness. | CG1, CG2, CG3, CG4, HCP1, HCP2, HCP3, HCP4, HCP6, HCP7, HCP9 |
| May trigger negative or aggressive emotional reactions. | CG1, HCP1, HCP4, HCP6, HCP7 |
| Reduced usability as dementia progresses. | CG2, CG3, HCP1, HCP7 |
| ***On the part of CGs & HCPs*** |  |
| Unpredictability of PWD reaction due to individual differences. | HCP2, HCP3, HCP7, HCP8, HCP9 |
| General uncertainty about added value of GRACE in comparison to other activities. | CG1, HCP1, HCP4 |
| Potentially requires additional effort. | HCP4, HCP7 |
| Time constraints in professional care settings. | HCP1, HCP4 |
| ***Technological challenges that are to be solved first*** |  |
| GRACE needs to recognize unclear or low-volume speech and be able to adapt to it (e.g. incoherent or inaudible conversations). | CG3, CG4, HCP3, HCP4, HCP8 |
| GRACE needs to adapt to unpredictable user behavior. | HCP8 |
| GRACE needs to accommodate vision and hearing impairments. | CG2, HCP4, HCP6 |
| ***Ethical concerns regarding the use of GRACE*** |  |
| Fear of replacing human caregivers with robotic alternatives. | CG4, HCP1, HCP4, HCP6, HCP8, HCP9 |
| Concern about the trade-off between personal data use and actual benefit. | CG1, CG3, CG4 |
| Concern about people with dementia forming emotional bonds with a robot. | HCP8 |
| Need for reliable deletion of personal data after use. | CG2 |
| Importance of obtaining informed consent for use of personal data. | HCP9 |
| Concerns about data security and risk of unauthorized access. | CG3, CG4 |
| Potential risk of deception of PWD through personalized interaction (e.g. when personalized topics are discussed or familiar sounds / voices are used). | HCP7, HCP9 |
| Concern about forced use and lack of user autonomy (e.g. GRACE should always be a voluntary option and never imposed on PWD). | HCP1, HCP2 |
| Fear of increased isolation due to reduced face-to-face contact. | HCP6 |
| Privacy concerns around passive listening in on background conversations in shared environments. | CG1 |
| Concern that GRACE may reduce joyful caregiver-PWD interactions. | HCP9 |
| Belief that all available tools should be tried if they help PWD. | HCP3 |

Some key quotes that highlighted a fear of contact with technology and low acceptance rate among PWD:

> *“My mom wasn’t very familiar with it [referring to a smartphone]. We tried to teach her, especially in the early stages, also to communicate via the internet, to get information via the internet. She didn’t want any of that”* (CG4)
>
> *“If they can’t understand or grasp something, or aren’t properly attuned to it, then it tends to make them aggressive.”* (HCP6)

Regarding ethical considerations, participants mentioned different aspects of data protection, where some could also weigh its risks against the benefits of using services like GRACE:

> *“It must be clearly regulated which data goes where (…) I have the feeling that I would have given a lot of information. In the end, it’s about the health and safety of a relative”* (CG3)
>
> “(..) how much does GRACE learn about my grandmother over time and how much is then stored in the background. But I don’t know, for some reason I’m less concerned about that, I’d be less concerned in that case. Well, I think the benefits would outweigh the problems” (CG4)

In summary, while respondents were generally open to GRACE, they emphasized that it is critical to address such challenges and ethical concerns to ensure that GRACE is used safely, beneficially, and voluntarily on an ongoing basis.

### Recommendations for the implementation and further development of GRACE

In addition to answering RQ2, respondents were also asked for recommendations on how to improve GRACE in the future, how to facilitate its introduction into practice and how to ensure user acceptance among all stakeholders involved. Table 7 summarizes feedback obtained regarding strategies for improving acceptance among PWD, implementation and familiarization, as well as integration into the workflows of CGs and HCPs.

**Table 7.** Recommendations for successful implementation and acceptance of GRACE as reported by respondents.

| Feedback | Mentioned by |
| --- | --- |
| **Strategies to Improve PWD Acceptance** |  |
| Encouraging PWD engagement through more motivation and praise. | CG1, HCP4 |
| Increasing acceptance through collaborative use of GRACE. | CG1, CG4, HCP2, HCP3, HCP6, HCP9 |
| Ensuring GRACE interactions are fun and entertaining. | CG1, CG2, CG4, HCP1 |
| Fostering positive early experiences with GRACE. | CG1 |
| Sparking interest by positioning GRACE as a personalized “friend”. | CG2, HCP3 |
| **Initial Implementation and Familiarization** |  |
| Boosting engagement through joint trial sessions with caregivers. | HCP1, HCP2, HCP6, HCP7, HCP9 |
| Improving acceptance through repeated exposure. | HCP1, HCP2, HCP3 |
| Motivating use by helping PWD recognize personal benefits. | CG1, HCP7 |
| Introducing GRACE with the help of younger caregivers (e.g., grandchildren). | HCP9 |
| **Integration into Workflows and Ease of Use** |  |
| Making integration easier through collaborative use. | CG1 |
| Providing enough familiarization time before full adoption to ease the learning curve. | CG4, HCP1, HCP3 |
| Facilitating adoption by helping caregivers understand GRACE's functions. | HCP1, HCP8 |
| Identifying the right physical placement for GRACE. | HCP6, HCP7 |
| Ensuring GRACE is intuitive and easy to use (e.g. 1-2 buttons at max.). | HCP1, HCP7 |
| **General Implementation and Integration Advice** |  |
| Enhance human-likeness through expressive facial features and natural interaction. | CG1, CG2, HCP1, HCP2, HCP7 |
| Ensure GRACE is intuitive, comes with a manual, and requires minimal setup. | CG4, HCP1, HCP8, HCP9 |
| Test GRACE in real-world settings with PWD before finalizing features. | HCP7 |
| Place GRACE at eye level in a consistent and visible spot to support interaction. | HCP2, HCP4 |
| Simplify choices and reduce decision-making demands for PWD. | CG1, HCP1, HCP2 |

For successful initial implementation, two critical factors emerged: joint use with PWD and thorough preparation of CGs and HCPs. The concept of familiarization was paramount, emphasizing that PWD, CGs as well as HCPs need to gradually acclimate to GRACE’s presence and interaction. This process involves trying GRACE together with PWD multiple times and doing so in an empathetic manner. Simultaneously, comprehensive introductions and training for CGs and HCPs on GRACE’s functions and benefits were named as important ingredients for a successful initial implementation. This could be promoted, for example, by making user manuals available and ensuring that setup is very simple and intuitive. In addition, to significantly boost PWD acceptance and motivation to use GRACE, respondents considered shared use with CGs and HCPs as crucial, as this could foster an enjoyable and engaging experience. Furthermore, they expressed that incorporating fun elements and praise into GRACE’s interactions could increase long-term engagement. Finally, making GRACE more human-like through natural facial expressions and speech patterns was also suggested. However, there were also some comments that viewed such humanization as rather negative, as it could potentially cause confusion among PWD.

To answer RQ3 (functional optimization), respondents provided several recommendations for improving and expanding GRACE’s general and technical functionalities (see Table 8).

**Table 8.** Recommendations for enhancing GRACE’s technical and general functional domains.

| Feedback | Mentioned by |
| --- | --- |
| **Technical Improvement** |  |
| Adding a screen or visual elements (e.g., for showing pictures). | CG2, CG3, HCP4 |
| Including headphone support for clearer audio. | HCP4 |
| Enabling connection to smart-home features (e.g., lighting control). | CG2 |
| Supporting behavior-based interaction for personalized responses. | CG2 |
| Making GRACE wireless to improve mobility and ease of use. | HCP7 |
| Ensuring GRACE is easy and intuitive to charge. | HCP4 |
| **User Experience & Interface Improvement** |  |
| Avoiding distress caused by unexpected movement (e.g., sensors). | HCP1 |
| Offering customization options for GRACE's look and feel. | CG2, HCP1, HCP 7, HCP8 |
| Providing different physical form options (e.g., radio, TV, puppet). | CG2, HCP1 |
| Keeping GRACE's interactions very simple and straightforward. | HCP1, HCP2 |
| Making interactions more friendly, motivating, and engaging. | CG1, CG2, CG4, HCP2, HCP3, HCP4 |
| Focusing on helpful content rather than excessive compliments. | CG2 |
| Simplifying physical controls (1-2 buttons max.) and ensuring clear on/off feedback. | HCP2, HCP4, HCP7, HCP9 |
| Using slower, simpler language in GRACE's spoken interactions. | HCP1, HCP2, HCP7, HCP8, HCP9 |
| ***Functional improvement recommendations for supporting PWD*** |  |
| Improving interaction through behavior-based tracking | CG1 |
| Adding more language and dialect options. | CG1, HCP4, HCP6, HCP7 |
| Personalizing interaction through user-specific preferences and topics. Potentially by using generative AI. | CG1, CG2, HCP1, HCP2, HCP3, HCP7, HCP8, HCP9 |
| Providing a calling function for emergencies. | CG1, CG2, CG3, CG4, HCP6, HCP8 |
| Helping maintain contact when phone PIN is forgotten. | CG3 |
| Offering mnemonic aids and reminder functions. | CG1, CG2, CG3, HCP3, HCP6 |
| Playing music tailored to the PWD's preferences. | HCP1, HCP2, HCP3, HCP7 |
| ***Functional improvement recommendations for supporting CGs & HCPs*** |  |
| Providing entertainment and conversation support for PWD. | HCP2, HCP4, HCP9 |
| Including a translation function for multilingual communication. | HCP6 |
| Supporting caregivers with task recommendations and checklists. | CG2, CG3 |
| Integrating a medication dispenser for easier daily care. | CG2 |
| Enabling early-stage disease diagnostics. | CG2, CG3, HCP9 |
| Supporting disease progression tracking. | CG1, CG2, CG3, HCP9 |
| Implementing intelligent emergency notifications. | CG2, HCP6, HCP8 |
| Tracking and monitoring PWD location and daily activities. | CG1, CG2, CG3, HCP8 |

The interviews also yielded numerous suggestions for new exercises in the areas of cognition, physical activity, creativity, and social interaction (see Figure 1), with open-ended questions and discussions about personal memories emerging as a particularly strong desire, underscoring the desire for more natural conversation opportunities.

**Figure 1.**
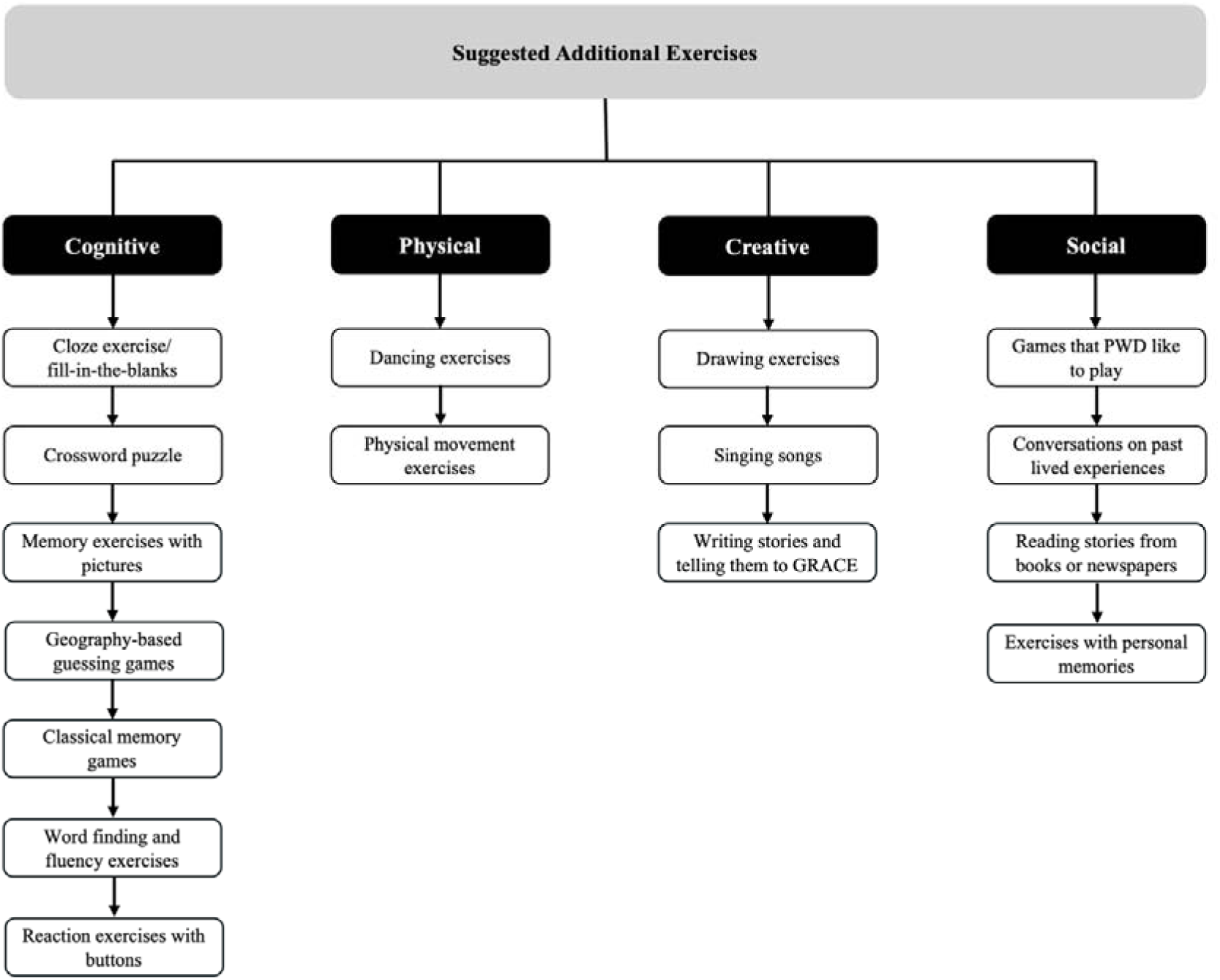
Additional activities suggested by respondents for GRACE, categorized by cognitive, physical, creative and social domains.

Beyond its current functionalities, respondents identified several additional potential applications for GRACE within dementia care that have been summarized in Table 9 (also contributing to RQ3).

**Table 9.**
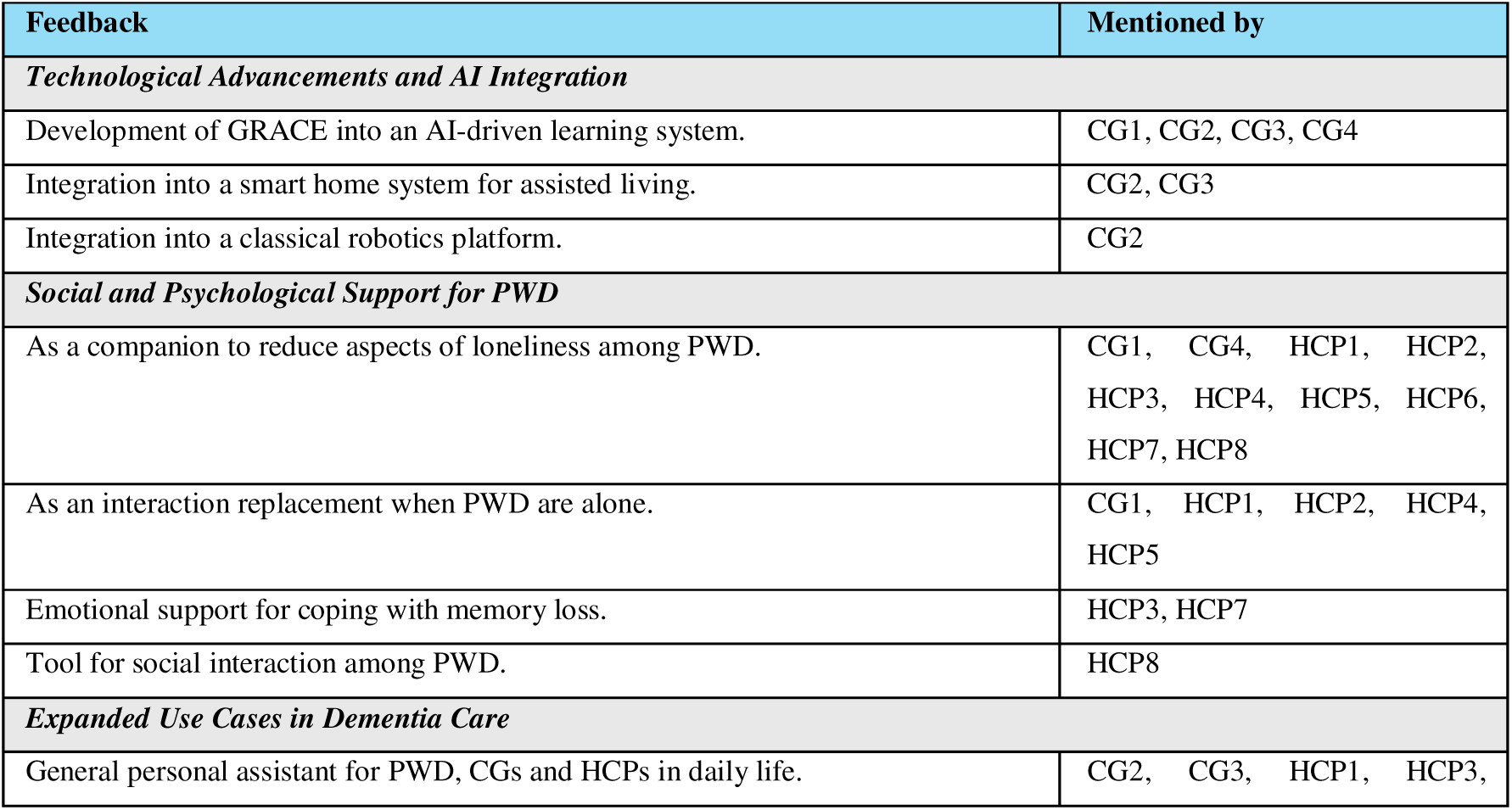
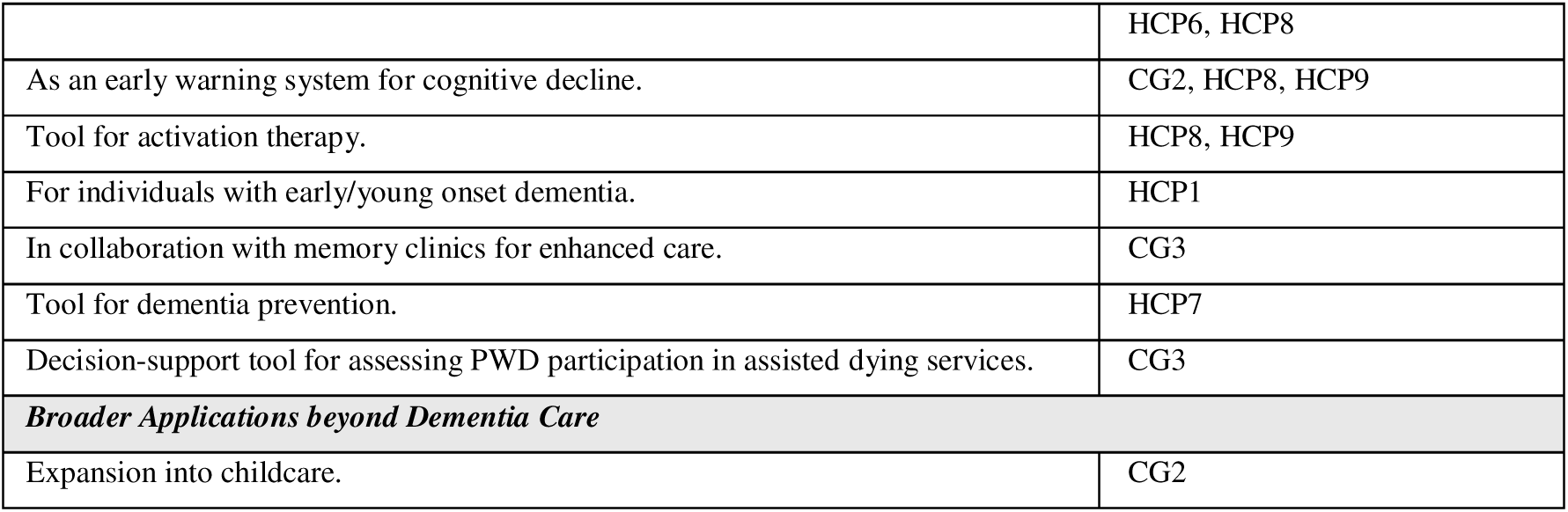
Potential future areas of application for GRACE.

The most frequently suggested broader role for GRACE was as a companion to reduce loneliness among PWD (CG1, CG4, HCP1, HCP2, HCP3, HCP4, HCP5, HCP6, HCP7, HCP8), and also as a general personal assistant for PWD, CGs and HCPs in their daily routines (CG2, CG3, HCP1, HCP3, HCP6, HCP8). Examples mentioned included assistance with household tasks, facilitating communication and coordination among HCPs, monitoring PWD’s condition, and aiding with various other daily responsibilities. Respondents also proposed that GRACE could include more AI-supported, personalized content and features (CG1, CG2, CG3, CG4), where it may also be possible to adapt to the PWD’s stage of dementia, potentially empowering PWD to remain in their homes longer (GC2, GC3, HCP1, HCP3, HCP6, HCP8), thereby enhancing their independence. Further suggested applications included utilizing GRACE as an early warning system for cognitive decline prior to a dementia diagnosis (CG2, HCP8, HCP9), and as a tool for activation therapies in clinical environments (HCP8, HCP9).

## Discussion

This qualitative evaluation of the voice assistant GRACE among HCPs and CGs provides both practical insights and guidance for future development for assistive technologies in this field of application, thus significantly contributing to filling the current gap in well-founded empirical research on how such technologies are perceived by those directly involved in dementia care and how they can be meaningfully integrated into care practice.

Overall, respondents were excited about GRACE’s potential to improve daily interactions with PWD and reduce the burden on HCPs and CGs at the same time, confirming the potential to ease caregiving responsibilities as described by Scheider et al. [4] if designed and implemented correctly and efficiently. Respondents particularly appreciated the memory exercise and the onboarding section. The former mainly for its potential to stimulate PWD cognitively and its thematic relevance to everyday life (e.g., shopping list) and the latter for its potential to build initial trust and make the start of interaction exciting and enjoyable. Despite this positive feedback, respondents also pointed out a variety of challenges and barriers to practical implementation. For example, the oftentimes rather low level of digital literacy and hence the possible skepticism toward talking to “robots” instead of people were frequently cited as challenges that could complicate initial implementation. In addition, the current rather harsh nature of GRACE’s responses to errors was criticized, as this could, in the view of the respondents, make PWD feel tested and thus create a feeling of discomfort, which in turn could have a negative impact on acceptance and user motivation. In addition, minor technical challenges were mentioned that need to be addressed when GRACE is used in practice. These include, for example, the targeted handling of fragmented, incomprehensible, slow, or inaudible speech patterns on the part of PDWs to avoid interruptions in conversation or frustrating interaction patterns. Additionally, from the perspective of HCPs and CGs, two challenges for successful implementation were highlighted. First, GRACE must be able to be seamlessly integrated into existing care processes without creating significant additional effort or complexity. Second, the opportunity costs incurred by using GRACE instead of human activities must be offset by clear added value, for example by bridging unstructured periods when caregivers are unavailable of busy with another task.

Participants also raised a set of ethical considerations that should be carefully examined. While most agreed that the benefits of greater inclusion and independence justify data sharing, they emphasized the need for transparent consent procedures, strict data deletion policies, and safeguards against excessive reliance on the system at the expense of human contact. Furthermore, concerns were raised that PWD could develop an emotional attachment to GRACE, which could further increase their social isolation. These observations are consistent with broader discussions on the ethics of digital health, which emphasize the importance of prioritizing human relationships and using technology only as a support rather than a replacement.

These findings both confirm and complement previous research on digital assistive technologies in dementia care. In line with Schneider et al. [4] our respondents saw clear potential for the voice assistant GRACE to provide cognitive stimulation and support everyday routines of PWD, CGs and HCPs. This observation also correlates with findings from further previous research [2, 4, 10, 19–21]. Similarly, multiple prior research [1–2, 10, 20, 22–24] emphasized the importance of user-centered adoption of such tools, which is reflected here in recommendations for thematic and conversational personalization, the described need for joint use in the beginning as well as the need of GRACE to provide a seamless implementation into existing workflows. Additionally, our findings show that low digital literacy among PWD and hence, a potential lack of user acceptance and motivation are the crucial expected barriers when it comes to the adoption of such assistive technology. This aligns with prior research identifying digital literacy and user acceptance as key challenges when implementing assistive technologies in dementia care [1, 10, 20, 24]. Furthermore, ethical concerns identified in this study also overlap with prior findings. Examples include the concerns expressed by respondents in this study regarding issues such as data protection and the risk of replacing people or further isolating them from social life through the use of such tools, which have already been highlighted in previous research projects [2, 10, 19, 25].

Based on these results, CGs and HCPs suggested several improvements to maximize GRACE’s usefulness and acceptance. Besides the joint use of GRACE (especially in early adoption stages), personalization was a recurring theme, ranging from tailoring conversation topics to the individual preferences of the PWD to adjusting voice tones and speaking speed to user needs. Interestingly, many envisioned GRACE evolving into a conversation partner able to provide open dialogue supported by generative Artificial Intelligence (AI), allowing the assistant to recall previous interactions and maintain small talk or memory activities in a more natural and human-like way. Respondents envisioned that GRACE could develop into a “companion” that could engage in free conversations with PWD, CGs and HCPs, provide reminders, help to combat loneliness on the part of the PWD, help facilitate coordination between caregivers, or proactively check in with PWD throughout the day. From a more professional point of view, CGs and HCPs also proposed additional support features to help caregivers with supporting PWD. Recommended features included appointment reminders, emergency call functions, dementia stage monitoring and the provision of practical advice from GRACE to CGs and HCPs. Many of these recommendations are also consistent with the findings of previous research projects that examined similar tools. For example, with Balasubramanian [19], who identified the potential of such tools to combat loneliness, with Bradford et al. [25], who investigated the provision of support functions for PWD, CGs, and HCPs, and with findings from other researchers who investigated the use of reminder functions for PWD, CGs, and HCPs [2, 10, 20, 24]. Finally, the results of this study also provide a glimpse into the future, as respondents suggested potential future areas of use for GRACE.

It can thus be said that the findings of this study both confirm earlier research results with empirical data whilst simultaneously adding to them with greater thematic depth. By systematically collecting the opinions of HCPs and CGs, our study provides solid insights into how the potential of voice assistants is perceived in real-world dementia care. While previous work has largely outlined general use cases, our results indicate exactly which intervention components are most valued in practice, which specific barriers need to be overcome, and how such a system can be integrated into the everyday lives of direct caregivers. We thus close an important knowledge gap regarding how tools such as GRACE can be developed responsibly, used ethically, and integrated sustainably into the everyday lives of PWD, CGs and HCPs. Furthermore, these findings provide concrete guidelines, prioritization criteria and insights on potential future areas of use that will guide the next iterations of GRACE’s features, interactions, and deployment strategies to ensure its continuous optimization and successful adoption into practice.

## Limitations

This qualitative study, which used inductive thematic analysis, collected feedback on the GRACE language assistance prototype from HCPs and CGs using qualitative interviews. Although the results are valuable for further investigation, some limitations should be considered when interpreting them.

First, the study included a relatively small sample consisting of eight HCPs and four CGs (five individuals). Although saturation was observed among the HCPs, it was less evident in the CG group. The diverse characteristics of the participants (e.g., technology affinity, age, experience) combined with a multifaceted sampling strategy may have led to selection biases that may limit the representativeness of our sample. For example, participants recruited through personal networks may have had a more positive opinion than those recruited through other channels.

Second, participants’ responses may have been influenced by several biases. Given the sensitivity of dementia care, social desirability bias is particularly likely to have an influence on the results. In addition, it is likely that memory biases also influenced the quality of the responses. This is partly because the CG respondents often reported on past experiences and partly because the interviews on GRACE were conducted exclusively based on a demonstration video sent out to them in advance. This could have influenced the depth and accuracy of the feedback provided and must therefore be considered when interpreting the results.

Third, although flexible, the semi-structured interview methodology led to variations in the topics discussed, which made it difficult to synthesize and compare participants’ responses. Inconsistent interview contexts, including different locations and occasional technical glitches, may have further influenced the participants’ answers.

Finally, limitations also arose from the analysis process. The inductive thematic analysis was conducted primarily by a single researcher, assisted by a second researcher, which may have introduced subjectivity and research-related biases in the interpretation and coding of the interviews. In addition, linguistic discrepancies arising from multiple translations from Swiss German to German for transcription and then to English for coding pose a risk of losing or distorting insights along the way.

## Conclusion

Overall, the findings from this study reveal that GRACE holds substantial promise as a supportive tool for PWD, CGs, and HCPs. Initial impressions indicated a high degree of openness and interest from both CGs and HCPs, along with constructive feedback on existing functionalities. The identified strengths of GRACE’s onboarding and the memory exercise, alongside detailed insights into areas for improvement in exercise design and responsiveness, provide a clear roadmap for iterative development. Importantly, the study identified significant potential barriers to adoption, primarily concerning PWD’s technological know-how, motivation to use, and the ethical implications surrounding human replacement, security and data privacy. The comprehensive recommendations for future enhancements, including technical refinements, user experience improvements, and expanded functionalities underscore the importance of user-centered and ethically conscious design. Addressing these study insights will be paramount in ensuring GRACE’s successful, beneficial, and safe integration into dementia care, solidifying its role as a valuable complementary tool rather than a replacement for human interaction.

## Data Availability

The datasets generated during and/or analyzed during the current study are available in the OSF repository at https://osf.io/qtxkc

## Acknowledgements

We would like to thank all the participants of the study, for generously providing their valuable time and perspectives to this study. We would also like to thank Ms. Vanessa Yarza Navarro-Schär for assistance during development of the interview guide.

## Author Contributions

FW: data curation, investigation, formal analysis, visualization, writing (original draft) & writing (review & editing). NBA: funding acquisition & writing (review & editing). TK: conceptualization, funding acquisition, methodology, supervision & writing (review & editing). RV: conceptualization, funding acquisition, methodology, project administration, supervision, validation, visualization, writing (original draft) & writing (review & editing).

## Statements and Declarations

### Ethical Considerations

On 17 September 2024, the Ethics Committee of the University of St. Gallen issued a letter exempting the study from a formal review. The exemption was granted because the interview study did not fall under the Swiss Federal Act on Research Involving Human Beings, did not expose participants to physical or psychological risk, nor involve vulnerable populations, or require collection of sensitive personal data.

### Consent to Participate

All participants were fully informed about the study and provided written informed consent prior to participation in the research study.

### Consent for Publication

All participants provided written informed consent for the publication and dissemination of research results obtained from their interview data in anonymized form.

### Data Availability

The datasets generated during and/or analyzed during the current study are available in the OSF repository, https://osf.io/qtxkc.

